# A Non-Invasive and Non-Contact Jugular Venous Pulse Measurement: A Feasibility Study

**DOI:** 10.1101/2024.06.04.24308313

**Authors:** Shatabdi Das, Girish Dwivedi, Hadi Afsharan, Omid Kavehei

## Abstract

The Jugular Venous Pulse (JVP) is a vital gauge of proper heart health, reflecting the venous pressure via the Jugular Vein observation. It offers crucial insights for discerning numerous cardiac and pulmonary conditions. Yet, its evaluation is often over-shadowed by the challenges in its process, especially in patients with neck obesity obstructing visibility. Although central venous catheterization provides an alternative, it is invasive and typically reserved for critical cases. Traditional JVP monitoring methods, both visual and via catheterization, present significant hurdles, limiting their frequent application despite their clinical significance. Therefore, there is a pressing need for a non-invasive, efficient JVP monitoring method accessible for home-based and hospitalized patients. Such a method could preempt numerous hospital admissions by offering early indicators. We introduce a non-invasive method using a frequency-modulated continuous wave (FMCW) radar for JVP estimation directly from the skin surface. Our signal processing technique involves an eigen beamforming method to enhance the signal-to-noise ratio for better estimation of JVP. By meticulously fine-tuning various parameters, we identified the optimal settings to enhance the JVP signal quality. In addition, we performed a detailed morphological analysis comparing the JVP and photoplethysmography signals. Our investigation effectively achieved signal localization within a Direction of Arrival (DoA) range from -20° to 20°. This initial study validates the effectiveness of using a 60 GHz far-field radar in measuring JVP.

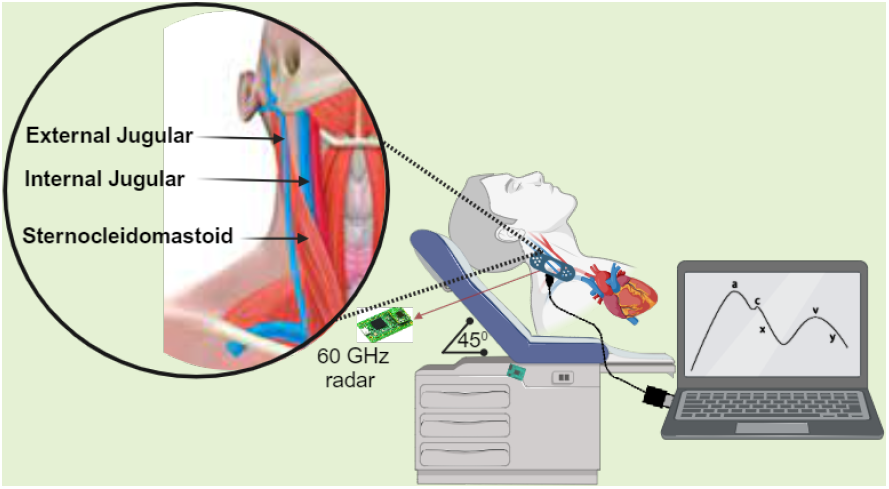

## I. Introduction

**H**EART Failure (HF), medically referred to as congestive heart failure, occurs when the heart fails to adequately pump the required amount of blood to meet the demand of the whole body. This situation can arise due to either the inability of the heart to fill up with an adequate blood volume or its weakened capacity to pump effectively. HF can potentially impair the functionality of vital organs, such as the brain, liver, or kidneys. In Australia, about 15% of hospitalizations are attributed to HF [1]. The yearly count of HF-related hospital stays surpasses 150,000 in Australia alone, resulting in an estimated cost exceeding $350 million due to the necessity of in-patient care following re-admissions [2]. This financial burden is anticipated to escalate due to the aging Australian population like many other countries worldwide.

Venous congestion is a critical factor in right heart failure, with symptoms developing gradually. This has increased emphasis on studying the role of the right ventricle in HF, with the Jugular Venous Pulse (JVP) being a crucial indicator of heart health [3]. JVP provides insight into heart and lung conditions by measuring venous pressure via the Internal Jugular Vein (IJV). The jugular veins, spanning from the head to the heart, consist of six vessels divided into three pairs. Illustrated in Fig. 1, the JVP waveform unfolds in five phases, highlighting a biphasic pattern. The ‘**a**’ wave signifies right atrial contraction, the ‘**x**’ descent follows, indicating atrial relaxation, and the ‘**c**’ wave emerges from right ventricular contraction. Subsequent ‘**x**’ descent marks the tricuspid valve’s downward movement, the ‘**v**’ wave denotes venous filling, and the ‘**y**’ descent represents rapid atrial to ventricular flow.

**Fig. 1:**
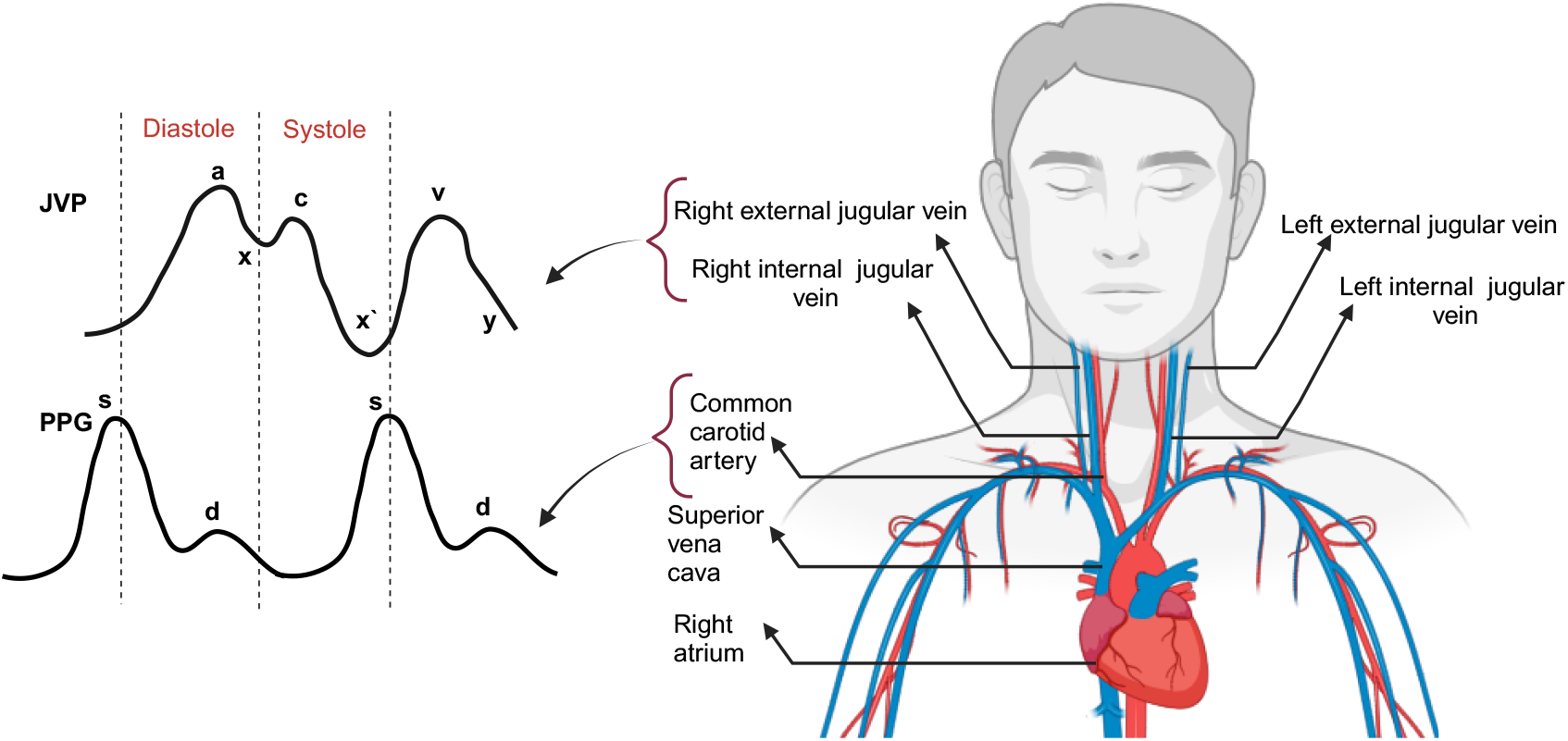
(a) Schematic representation of the superficial venous system of the inferior anterior area of the neck which connects the IJV, External Jugular Vein (EJV) with the right atrium (b) Interpretation of jugular venous pulse contour.

The ‘**a**’ wave in JVP, indicating atrial contraction, precedes the photoplethysmography (PPG) systolic peak, offering insights into cardiac timing. The ‘**x**’ descent in the JVP, representing atrial relaxation during ventricular systole, parallels the rising phase of PPG. The ‘**c**’ wave, related to the tricuspid valve movement, coincides with the PPG peak but lacks a direct PPG analog. The v-wave, associated with venous filling, aligns with the dicrotic notch of PPG, marking the aortic valve closure. The ‘**y**’ descent, indicating atrial emptying into the ventricle, corresponds with the declining phase of the PPG, signaling the cycle end and the readiness for the next heartbeat.

Traditional JVP measurement techniques, such as central venous line catheterization, are invasive and limited mainly to critically ill patients in intensive care or coronary care settings due to the high risk of complications. This technique necessitates critical staffing and key technologies for signal transmission and recording. Therefore, clinicians require a reliable, non-invasive, and quick technique to monitor JVP. Also, accessibility (or ease of access) to both at-home and hospital patients is another important factor that plays a crucial role in the early detection of HF.

Studies have investigated plethysmography as a potential method for estimating JVP; however, the dependency of PPG technology on the optical properties of skin and tissue introduces significant calibration challenges, adding complexity to its application by restricting patients from moving or changing positions [4], [5]. On the other hand, RF sensing techniques have emerged as a promising alternative, thanks to their efficacy in tracking physiological changes. Radar technologies have been explored to monitor vital signs, including blood pressure and heart rate, with specific investigations into 24 GHz microwave radar [6] and 900 MHz near-field radar [3] for JVP measurement. The 900 MHz band, while offering deep penetration, may not deliver the spatial resolution needed for detailed JVP waveform analysis. Our preference for a 60 GHz radar sensor over a 24 GHz one stems from its ability to offer greater resolution due to its higher frequency, enhancing its sensitivity to minor, surface-level movements. Such improved resolution proves advantageous in precisely delineating the subtle nuances of JVP waveforms. Specifically, we have chosen Infineon Technologies AG’s BGT60TR13C radar sensor since it provides a sophisticated integrated antenna design that minimizes susceptibility to interference and, most importantly, is flexible towards tailoring different parameters according to our needs.

We also utilized ZMT Zurich MedTech AG’s Sim4Life electromagnetic simulator to explore the impact of radio-frequency electromagnetic fields (RF-EMF) on the neck region, specifically examining the effectiveness of higher-frequency radar in estimating the JVP from the skin surface. Sim4Life studies have revealed that the external jugular vein is close to the skin surface. This proximity allows higher-frequency radar to concentrate its effects in this area, minimizing the dispersion or scattering of electromagnetic energy through the skin layers. Consequently, this radar technology is highly effective for applications that require precise targeting just beneath the skin surface.

For the first time, this study introduces the use of Frequency Modulated Continuous Wave (FMCW) radar technology for the non-invasive extraction of the JVP. FMCW radar technology, with its afford-ability and sensitivity, offers a promising alternative to patients at risk of hospital readmission, including those in urban, rural, and remote areas of Australia. The self-administered, non-invasive JVP measurement could significantly reduce hospital readmission by facilitating early intervention.

## II. Related Work

Numerous research studies have explored various techniques for assessing JVP, including invasive, non-invasive, contact, and non-contact methods. In the following sections, we will describe these findings.

### A. Invasive Method

The most common and widely available method for measuring JVP is cannulation, an invasive technique considered the gold standard. In this procedure, an experienced anesthesiologist inserts venous catheters into the right internal jugular or subclavian vein (SVC). These catheters are then connected to a multi-modular monitor via a pressure transducer. After zeroing the transducer to the heart level, the mean venous pressure (mm Hg) is displayed in real-time on the monitor and recorded as invasive venous pressure [7].

Invasive methods can lead to various complications related to vascular access and typically require trained and skilled physicians to perform.

### B. Non-invasive Contact Method

The non-invasive are classified as intermittent and continuous. Developing a continuous monitoring technique for JVP is a primary concern for the future of remote heart patient monitoring. Recent research has defined several continuous, non-invasive JVP measurement methods based on pressure sensors, ultrasound, and optical PPG methods.

Irene García-López et al. [4] proposed an approach where reflectance contact PPG is used to sense JVP and obtain its physiological parameters. The PPG sensor was placed on the upper chest to trace the JVP. The potential of this method lies in its wearability and use of a single point of contact, making it a promising candidate for a wearable device. However, the inclusion of an infrared camera increases the cost, and setting up the device on the appropriate venous system can be complex.

Another non-invasive technique used accelerometer sensors to measure the vibrations of the jugular vein and the carotid artery [8]. This method can monitor the skin motion caused by blood volume changes in the IJV. Since the method relies solely on skin motion to translate the JVP waveform, patient movement can scramble the signal. It is also challenging to use with obese patients.

Conroy and colleagues proposed a wearable near-field RF sensor affixed with a neck collar on the clavicle over the IJV to enable noninvasive JVP sensing [3]. While this method is more convenient compared to others, the precise quantification of CVP extracted from JVP is not possible due to its single-point measurement.

### C. Non-invasive Non-contact Method

Imaging-based PPG, where the jugular waveform can be consistently observed non-invasively, is a non-contact JVP assessment approach suggested by Robert Amelard et al. [9]. This technique offers the advantage of ensuring the utmost comfort among all JVP sensing methods in a clinical environment where subjects are relatively immobile confined to a bed. However, it imposes notable limitations on patient positioning. Patients must maintain a consistent orientation towards the camera and hold the same posture throughout the recording, introducing complexities for continuous or frequent long-term monitoring [3].

The most well-established alternative approach to sensing JVP involves ultrasound imaging of the IJV structure [10] [11] [12]. Bedside ultrasound of the IJV, performed by emergency physicians, provides immediate, important information that cannot be obtained without invasive catheters. However, Wang et al., [13] claimed that their study was carried out on a small number of patients and was not validated against the gold standard method. Nonetheless, ultrasound utilization involves significant costs, including the need for specialized equipment and trained operators. This restricts its applicability to clinical settings and short-term recordings, preventing its widespread use for continuous or ambulatory JVP sensing in the long term.

A recent experimental study developed a prototype non-contact system that employs microwave radar (24 GHz, 7 mW) for JVP measurement [6]. The main challenge lies in the qualitative nature of pulse description and the variations in interpretation provided by a medical professional of the inherent conditions. Consequently, mastering pulse palpation techniques becomes arduous, and the diversity in these techniques contributes to disparities in pulse analysis.

In summary, it can be understood that JVP effectively reflects changes in right atrial pressure due to its unique valve arrangement, distinguishing it from other veins. However, its clinical application is limited due to the challenging nature of the measurements. JVP normal values range between 4– 6 mm Hg. Despite JVP details being present in the peaks and troughs of heartbeats, their small magnitudes often lead to oversight, rendering JVP an inefficient index. Palpation-based measurement involves vein compression,making pulse detection difficult. Hence, monitoring JVP requires high sensitivity to capture these minute signals without obstructing the vein.

Utilizing microwave radar sensors has recently gained popularity over other methods for monitoring vital signs. This method enables non-invasive tracking of vital signs, eliminating direct body contact. It can detect minuscule body surface motions resulting from cardiac and respiratory activities. The antenna settings allow for precise control over the measurement area. This technique has been practically employed, such as with a ceiling-mounted microwave radar placed approximately two meters above the subject [14]. This radar accurately monitors respiratory rates even through dense bedding. Furthermore, successfulmonitoring of Heart Rate Variability (HRV) has been demonstrated.

Additional research has been conducted on measuring blood pressure and other vital signs using continuous wave radar [15], FMCW radar [16] [17] [18]. Given that the characteristics of radar techniques align with the requirements for JVP measurements, there is potential for application in this context. Such utilization could alleviate the need to subject patients to significant physiological stress.

Using this technology can also give easy access and affordability for HF patients in remote areas. Enhancing the efficiency and performance of the technology can benefit clinicians and specialists involved in HF patient care, potentially influencing decisions regarding re-hospitalization or readmission. Hence, this current study aims to validate the efficacy of a non-invasive approach employing FMCW radar for JVP measurement. In this paper, we develop a system based on FMCW radar for JVP measurement. Furthermore, the performance of the system is assessed through experimental data. Additionally, we will explore the potential of the method for future utilization.

## III. Concept of FMCW RF system

Unlike a CW radar, an FMCW signal offers the added benefit of achieving range resolution due to its varying emitted frequency [16]. The FMCW system creates a linear signal that reflects as an echo upon encountering an object. This echoed signal is captured, combined with the transmitted signal, filtered, and converted into an Intermediate Frequency (IF) signal before being displayed. A simple representation of the FMCW RF system is shown in Fig. 2.

**Fig. 2:**
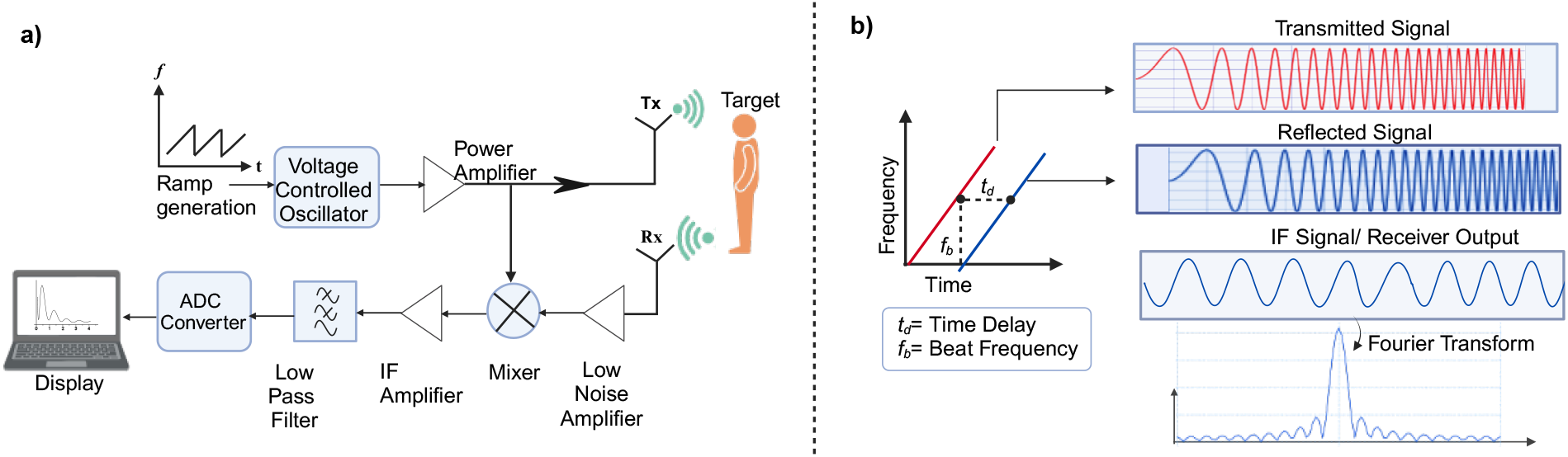
(a) A typical FMCW system with one transmitter and one receiver outlines the linear saw tooth ramp generation and the necessary modules for data processing. The received signal is amplified and low-pass filtered before being converted to digital representation. (b) The basic working principle of an FMCW radar system highlights transmitting a frequency-modulated signal and mixing it with the incoming signal at the receiver. This action generates an IF signal, represented in the figure as a sinusoidal waveform corresponding to the beat frequency.

The frequency of this IF signal corresponds to the beat frequency, which is utilized to determine the target range. The IF signal contains sinusoids with varying frequencies corresponding to the target ranges in multiple-target scenarios. The In-Phase (I) and Quadrature (Q) data obtained from an FMCW radar captures the frequency difference resulting from the delay between the transmitted and reflected signals, considering that the transmitted signal’s frequency varies over time. When the transmitted frequency-modulated signal interacts with a target, the received signal is delayed in time compared to the transmitted signal. The transmitted signal can be represented as;

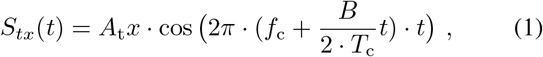

where, *A*_tx_ is the amplitude of the transmitted signal, *f*_c_ is the start frequency (carrier frequency) of the chirp, *B* is the bandwidth of the chirp, *T*_c_ is the chirp duration and *t* is time.

A chirp is characterized by the start frequency (*f*_c_), Bandwidth (*B*) and duration (*T*_c_). The time it takes for the chirp to reach the upper frequency from the lower frequency is called the chirp duration (*T*_c_). The slope of a chirp defines the rate at which the chirp ramps up. Bandwidth *B* is the difference between the upper and lower frequencies.

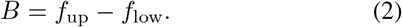

The received signal, which is a time-delayed version of the transmitted signal due to the round-trip travel time to and from the target(*t*_d_), can be represented as:

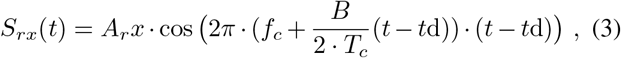

where, *A*_rx_ is the amplitude of the transmitted signal, *t*_d_ is the time delay of the received signal.

Fig. 2(b) demonstrates the computation of the beat frequency through a Fast Fourier Transform over the IF signal, enabling the estimation of the target range. The figure shows the linearly increasing frequency of the transmitted and reflected signals. Since the speed of electromagnetic waves is constant, this time delay can be used to calculate the distance to the target using the formula;

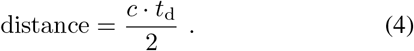

The beat frequency (*f*_b_) is the difference between the transmitted signal frequency and the frequency of the received signal after it has been reflected off a target. Beat frequency is directly proportional to the time delay and the distance to the target. In FMCW radar, this beat frequency is used to determine the range of the target using the formula;

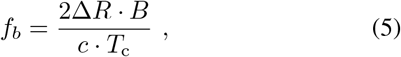

where, Δ*R* is the range resolution, *B* is the bandwidth of chirp, *c* is the speed of light and *T*_c_ is the chirp duration.

The system can determine the target distance in relation to its position and utilize the reflected signal solely from a designated range bin or a range span encompassing multiple bins. The maximum distance can be obtained by,

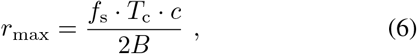

where, *f*_s_ is the sampling rate and *T*_c_ is chirp duration as mentioned previously.

FMCW radar operates best near the target area [17]. Thus, the FMCW system is believed to return only the most immediate range skin displacement, while we chose to work with FMCW for JVP measurement.

## IV. System Design

### A. Simulation Setup

The finite difference time domain (FDTD)–based solver of the commercially available software Sim4Life (Zurich MedTech AG, Switzerland) was used for all dosimetric evaluations. The FDTD method is computationally efficient for studying the electromagnetic (EM) exposures of complex heterogeneous dielectric structures, such as anatomical models.

An MRI-based full-body voxel model, Duke V3.0, was obtained from a virtual population for the human body model. The model was segmented into approximately 80 anatomical body tissues and organs, with a resolution of 0.8 × 0.8 × 0.8 mm^3^ throughout the body. The height and weight of the model were 1.63 meters and 57.3 kg, respectively. Parameters, such as permittivity, permeability, and electric conductivity, were assigned to the tissues according to the IT’IS Database.

A phased array antenna Fig. 3(a) is used as an EM source, with the antenna array excited with an input power of 350 mW. For computational analysis, a limited region (head including neck) is simulated, represented by a white boundary in Fig. 3(b). The simulated region (neck) accurately represents the skin, fat layers, muscle, and blood vessels (jugular veins and carotid artery) as shown in Fig. 4.

**Fig. 3:**
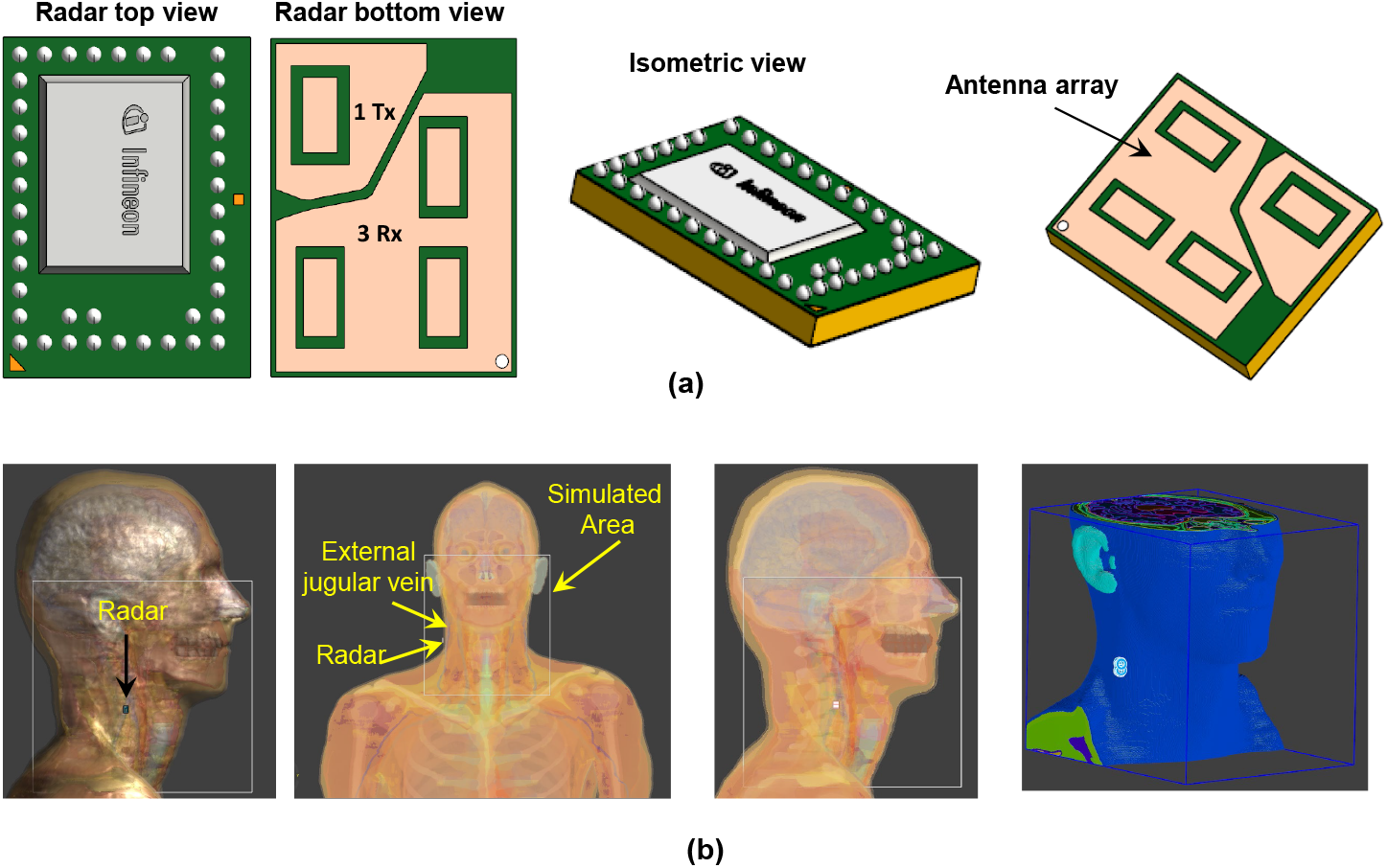
Overview of an FMCW radar simulation setup targeting the external jugular vein. The setup includes (a) isometric view of the radar system with phased array antenna-integrated RADAR model featuring 1 transmitter (Tx) and 3 receivers (Rx), and (b) simulation setup and radar positioning relative to the simulated area, focusing on the electromagnetic effects and SAR analysis in Sim4Life utilizing Duke model.

**Fig. 4:**
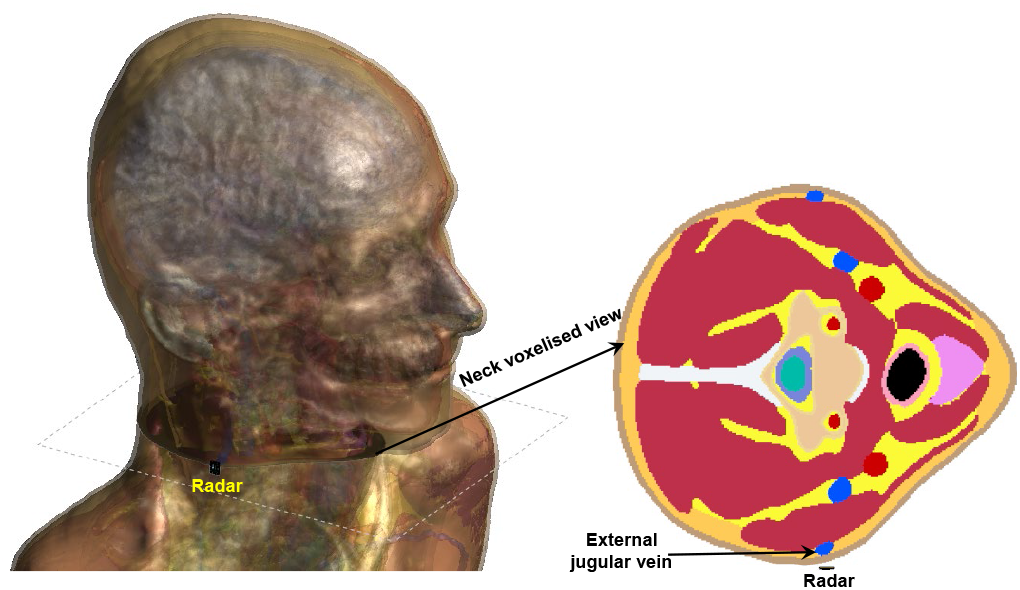
Realistic human model (Duke) accurately represents the skin, fat, muscles, and various other tissues.

In EM simulations, specific absorption rate (SAR) and power density (PD) are critical parameters for understanding the interaction of electromagnetic fields with biological tissues. In simulations conducted at 60 GHz, the wavelength of the electromagnetic wave is notably short, measuring approximately 5 mm in air and further reduced in biological tissues due to their elevated permittivity. Consequently, accurately modeling the electromagnetic fields at this frequency necessitates an excellent spatial resolution for the simulation mesh. This level of detail significantly increases the computational requirements, including memory and processing power, needed for precise simulations. Therefore, we have considered three different frequencies (900 MHz, 5 GHz, and up to a maximum of 35 GHz) for the SAR and PD analysis of the neck region. The simulation results are discussed in Sec. VI.

### B. Radar Board Specification

The radar selected for acquiring data in this study is off-the-shelf, relatively low-cost Infineon Technologies AG’s 60 GHz FMCW radar chipset (BGT60TR13C), with one transmit (*T*_x_) and three receive (*R*_x_) antennas. The receive antennas are positioned at right angles to each other. The radar sensor is in a laminated package with dimensions of 6.5 × 5.0 × 0.9 mm^3^. It operates within the 58 to 63 GHz range and offers customization options for samples per chirp (*N*_s_), chirps per frame (*N*_c_), and chirp duration (*T*_c_). The gain of each receive and transmit antenna is 10 (dBi) and 6 (dBi), respectively. This chipset creates precise linear frequency modulations.

The high sensitivity of this sensor allows for sub-millimeter motion detection. Generally, as the emitted frequency of a radar increases, its ability to detect subtle movements improves. Higher frequencies have shorter wavelengths, which amplify the signal’s phase with repeated motions [17]–[19]. Therefore, we selected to work with a 60 GHz system for measuring JVP, skin movement, or any displacements of very small magnitude.

### C. Radar Data Configuration

The radar data within a measurement frame is structured as a three-dimensional cube in the time domain Fig. 5(a). Fast time refers to the time scale within a single chirp associated with the samples per chirp. Slow time is the time scale across multiple chirps associated with chirps per frame. Both range and velocity information can be obtained by performing the Fast Fourier Transform (FFT) along the fast and slow times, respectively.

**Fig. 5:**
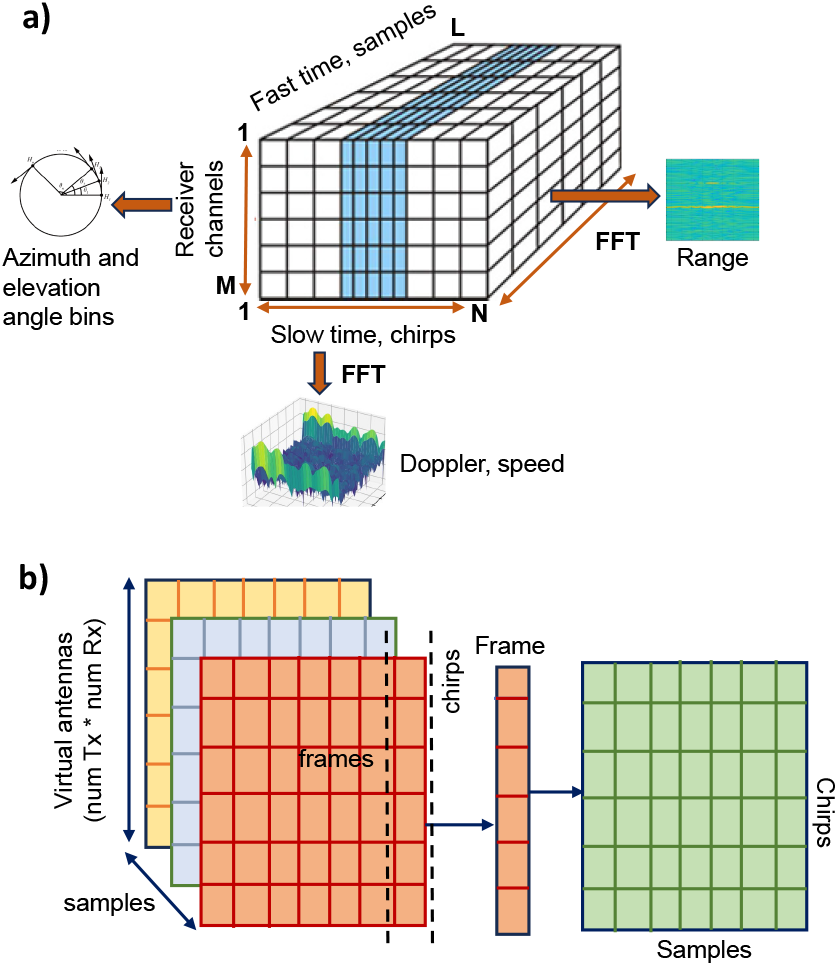
(a) Radar data cube representation. The data cube is structured along three axes: chirps (slow time), receiver channels, and samples, facilitating the processing of signals through multiple FFTs to determine range, Doppler speed, and angles in azimuth and elevation. (b) The frame structure of the data received by the radar sensor. Each frame comprises several chirps, and each chirp contains several samples.

Fig. 5(b) illustrates an example of raw radar data sample indices. Here, rows represent the number of receive (*R*_x_) antennas for each active transmit (*T*_x_) antenna available in the device, and columns indicate the number of chirps configured in a frame (*N*_c_), and slices indicate the number of samples per chirp (*N*_s_). Data from each transmit-receive antenna pair is treated as data from a single virtual antenna. The ordering of virtual receive antennas follows the (Tx,Rx) sequence based on the count of active transmit and receive antennas. When the digitized signal is formed into a frame, it assumes the structure of (*N*_Rx_ × *N*_*rmc*_ × *N*_s_), which is then utilized for signal processing. The number of samples per chirp indicates the fast time, whereas the number of chirps per second indicates the slow time.

### D. Experimental Setup

The radar was first updated to the latest firmware version v.2.55. For interfacing with the system, we utilized the latest version of the *Radar System Development Kit*, v.3.3.0, provided by Infineon Technologies AG, which is compatible with MATLAB (2023a, Math Works, Massachusetts, USA). Additionally, we designed a meshed cover with dimensions of 30 × 10.5 × 2 mm^3^ using Fusion 360 and 3D printed it by Creality, which is a biocompatible plastic in a laboratory environment as shown in Fig. 7(b).

Our trials across various distances have demonstrated the importance of correctly positioning the radar to achieve optimal signal strength. Proper positioning prevents saturation due to proximity and avoids losing details when the radar is too distant. Appropriate separation enhances signal processing efficiency and ensures precise tracking of physiological movements influenced by changes in the signal return time and frequency attributes.

To complete our setup, the next step was to identify the optimal neck location and adjust the subject’s position to facilitate the detection of the jugular vein. Venous return, cardiac output, and arterial and venous pressures are all affected by gravity. The ideal angle for measuring the jugular pulse falls between 30^°^ and 45^°^. Individuals with visible jugular distension do not require specific positioning. However, in healthy individuals, the jugular veins are usually not prominent when standing but may become distended to varying degrees in a lying position. Therefore, selecting the appropriate intermediate position allows for the observation of the peak of the column in the neck area between the collarbone and the jawbone. Thus, maintaining this inclination is crucial for accurately identifying JVP in healthy individuals and those with venous pressure conditions.

Now, to narrow down the area of the jugular vein on the neck, we considered the Small Intestine (SI) acupuncture points SI16 and SI17 as our reference. The Fig. 6 shows the position of SI16 and SI17 on the neck. SI16 and SI17 are acupuncture points near the jugular vein, yet neither directly overlaps with the vein due to their respective positions relative to the sternocleidomastoid (SCM) muscle [20]. SI16 is situated on the lateral aspect of the neck, posterior to the SCM muscle, and approximately level with the laryngeal prominence (Adam’s apple). Given its location posterior to the SCM, SI16 is somewhat lateral (to the side of) but near the internal jugular vein, which lies deep and medial to the SCM muscle.

**Fig. 6:**
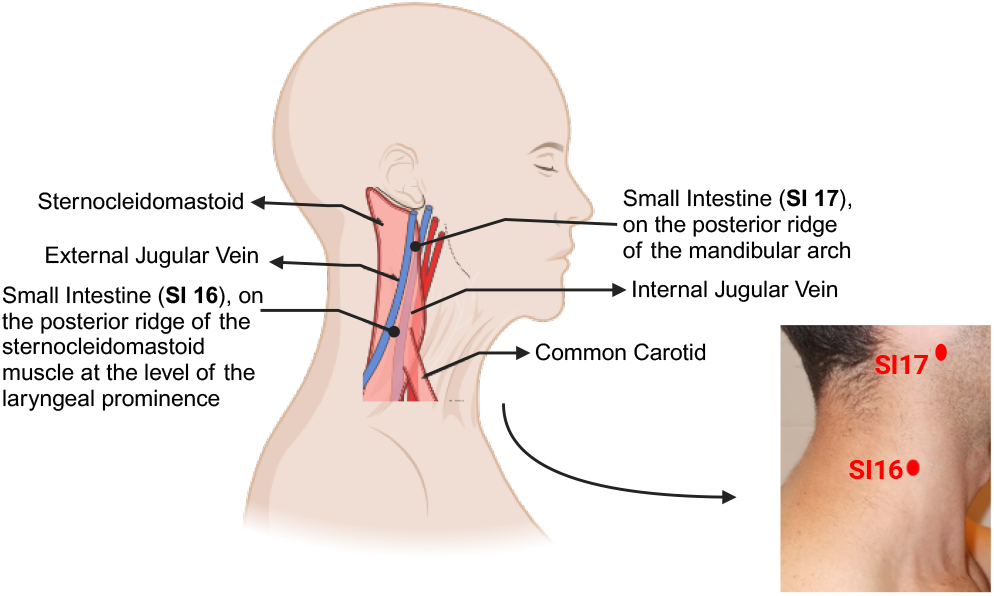
Using Acupoints (acupuncture points) SI16 and SI17 to identify jugular vein Locations. This diagram illustrates using the Small Intestine meridian acupoints SI16 and SI17 as anatomical references for locating the jugular veins. SI16 is positioned just posterior to the sternocleidomastoid muscle at the level of the laryngeal prominence, and SI17 is along the posterior ridge of the mandibular arch. Both points serve as key landmarks for identifying the paths of the external and internal jugular veins.

SI17 is located slightly differently in the depression between the SCM muscle and the mandible (jawbone) and anterior to the edge of the SCM muscle. This places SI17 closer to the path of the internal jugular vein as it runs deep and somewhat medial to the SCM. However, like SI16, SI17 is not directly over the vein.

After carefully considering both acupuncture points, we preferred SI16 as the most optimal choice for the radar placement. As a proof of concept and in adherence to ethical guidelines, this experiment was conducted on one healthy subject (female, aged 26 to 30 years, with a warm skin tone). The subject was positioned at varied angles of [0^°^, 30^°^, 45^°^, 90^°^] and the recording period was 30s each time. The device was set up for recording, and the data was collected on the host computer. Furthermore, to analyze the shape of the jugular venous waveform against the cardiac activity stages of systole and diastole, we gathered finger PPG data Fig. 7(a). The sampling rate of the single-channel PPG recording was 273.067 Hz, and the supply voltage was 5 V. Additionally, it is worth noting that multiple datasets were gathered across different setups of parameters (such as samples per chirp, chirps per frame, sampling frequency, and chirp duration) for experimentation and analysis. All the data were then processed and analyzed using MATLAB. This process aimed to identify the best parameters for accurately detecting the jugular venous waveform.

**Fig. 7:**
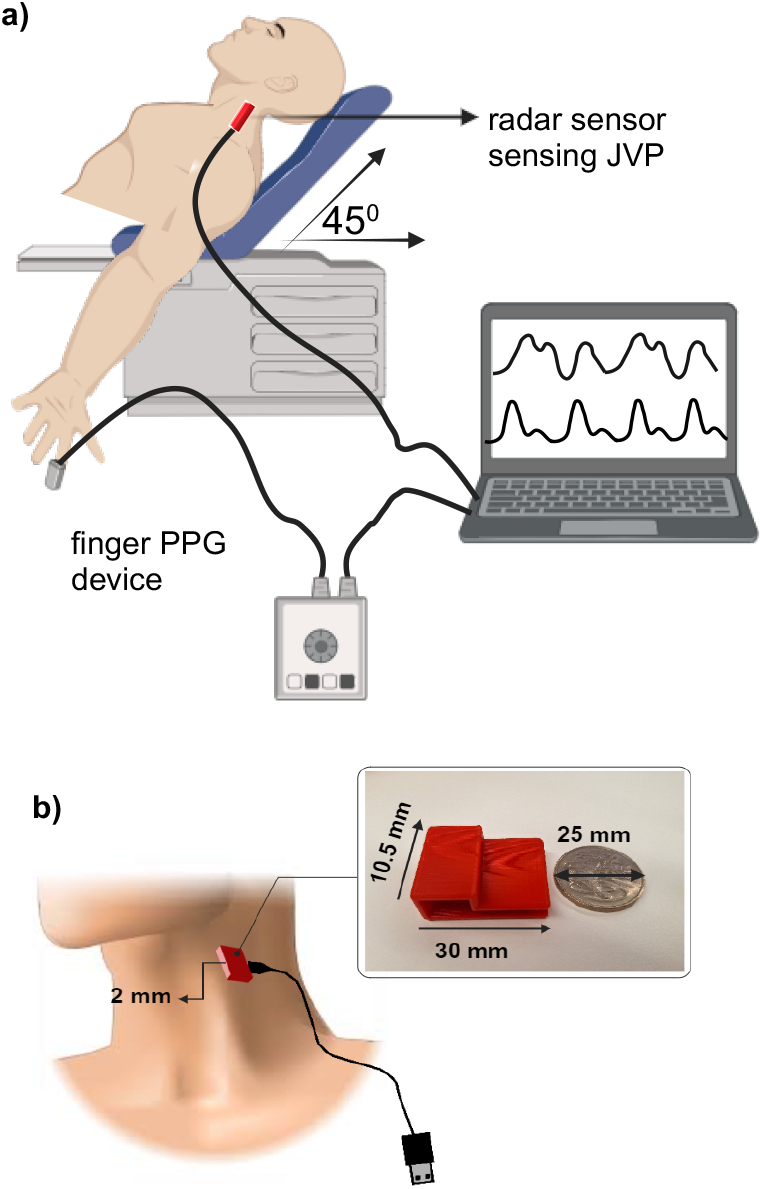
Data recording setup. (a) The radar sensor is placed near SI16 on the jugular vein for data collection, while the PPG data is collected using the finger PPG device. The subject is reclined with a sensor placed at the neck at an almost 45^°^ angle to monitor cardiovascular parameters. (b) The 3D-printed plastic case is nearly the size of a 1-dollar Australian coin.

(For further details regarding the specific device used in this study, including its configuration, interested readers are encouraged to contact the corresponding author^1^.)

## V. Methods of Data Analysis and Signal Processing

The workflow for the detection of the JVP is described in this section. The data cube contains the normalized ADC samples of the received signal. After obtaining the frame data (*N*_rx_ × *N*_c_ × *N*_s_), sampled at the beat frequency, is transformed into a complex range profile through the application of an FFT over the fast time dimension. Peaks within the FFT spectrum of the range indicate the presence of a target within the mmWave radar’s detection area. The overall data processing sequence is shown in Fig. 8.

**Fig. 8:**
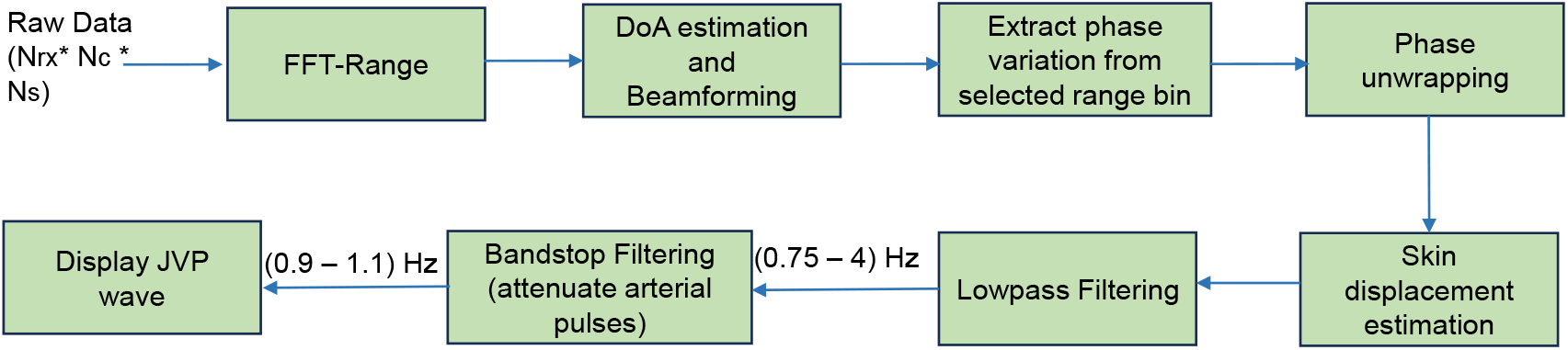
Schematic representation of the data processing sequence. After reshaping the raw data, an FFT is performed on the samples to obtain the range information. Upon computation of FFT, displacement is calculated from the phase variation, which is then used to extract the JVP.

JVP signals are venous pulsations reflecting changes in the right atrium’s pressure during the cardiac cycle. These changes are subtler and produce lower amplitude signals than the strong, rhythmic contractions of the heart or the more pronounced movements associated with breathing. Therefore, JVP, being more delicate than signals from breathing, might be masked within radar data. Spotting deviations in JVP signals also presents its own set of challenges. The lack of standardization in experimental frameworks across the field complicates comparing research findings. To tackle these issues, we introduced a simulator for vital signs, emphasizing JVP signal emulation through sound speaker vibration estimations. This allows a standardized environment to assess and benchmark algorithms tailored to JVP signal detection.

As illustrated in Fig. 9, we created a simulation of a patient target using a speaker plate designed to replicate human skin positioned towards the radar system. Due to the speaker’s operational frequency range, we broadcast a synthesized audio signal (beat signal) comprising two tones, *f*_1_ = 50 Hz and *f*_2_ = 54 Hz of the form:

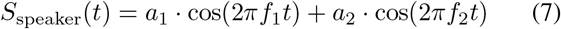

**Fig. 9:**
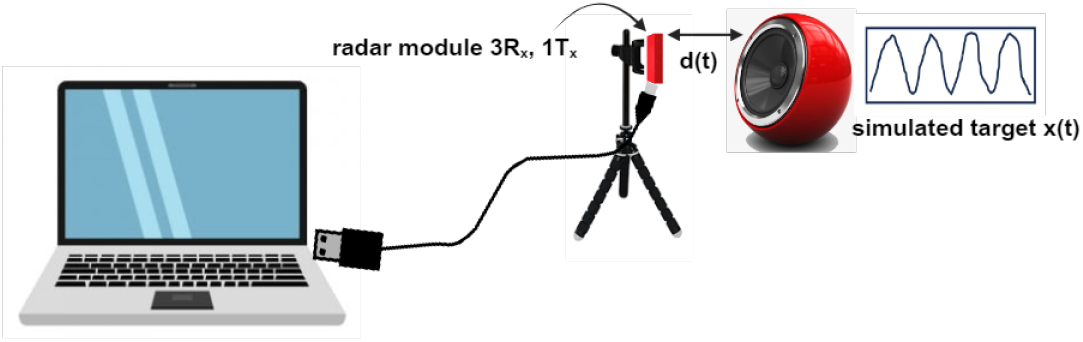
This setup features an experimental design replicating a patient target, with a speaker plate strategically oriented to face the radar system. The experiment involves the synthesis of an audio signal, meticulously composed of two specific frequencies, 50 Hz and 54 Hz, to evaluate the radar system’s response.

The genesis of this experiment was influenced by [21], where their setup was used for signal processing validation. The outcome of this experiment is discussed in Sec. VI. Below, we detail each step in the signal processing sequence, accompanied by the corresponding pseudo codes:

Step 1: Reshaping raw data vector into a matrix

- Size of matrix: (No of frames) *×* (size of frame)
- Size of frame: No of chirps per frame × 3 × No of samples per chirp

Step 2: Processing every antenna’s signal

- Applying FFT processing for every chirp pulse in each frame and to each antenna. (Optional windowing technique can be used before FFT for spectrum interpolation)
- Averaging FFT outputs to reduce the noise effect for each antenna signal and from each frame.

Step 3: Extracting IQ signal from each antenna

- From the averaged spectrum of raw data, each antenna detects the first spike as radar echo from the patient skin.
- Inverse Fourier transform (IFFT) of first spectrum bin to extract IQ complex signal for each antenna.

The procedure initiates with the acquisition of raw radar data. The raw data is subjected to an FFT per-chirp basis for each antenna. This is executed separately for antennas 1 and 3, as indicated by **(range_FFT_1)** and **(range_FFT_3)**. Following the FFT, the algorithm computes the average of the Fourier-transformed data across all frames for each antenna. This averaging process is crucial for noise reduction and signal clarity enhancement. Subsequently, the algorithm employs an inverse FFT (IFFT) on the average FFT data’s first bin to yield the final I/Q data for antennas 1 and 3. This step essentially reconstructs the time-domain signal from the frequency-domain data while focusing on the first bin, which indicates isolating the DC component or the specific frequency component of interest.

### Algorithm 1 I/Q Samples Extraction for Each Radar Antenna

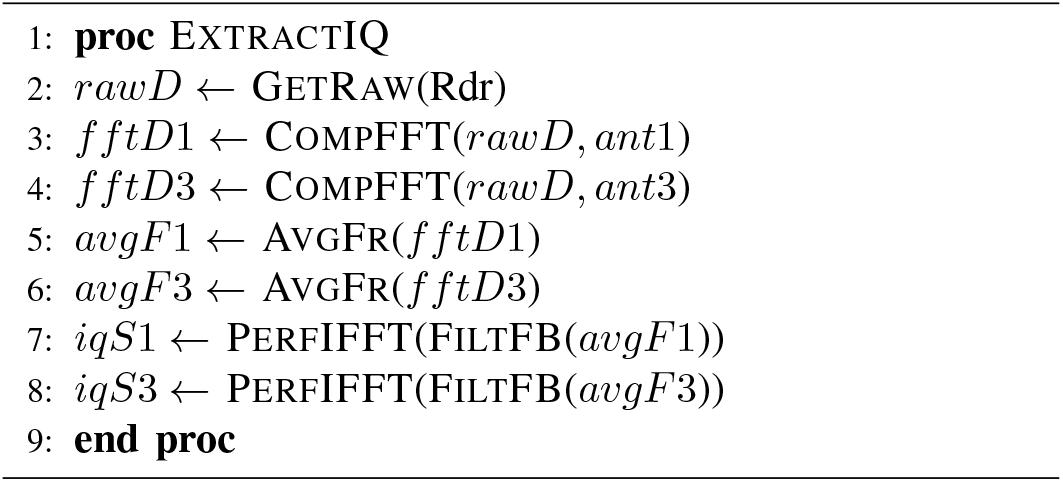

### A. Beamforming processing to enhance signal SNR

Referencing the I/Q data extraction process in [17], which applied the radar for monitoring blood pressure, our approach diverges in beamforming methodology. The eigenspace-based beamforming method (ESBM) used in their work encountered challenges with ill-conditioned matrices in our setup. Consequently, we have implemented the eigen-beamforming method, an adaptive technique that capitalizes on the eigenstructure of the signal to adjust the antenna array weights optimally. This ensures a stable and effective beamforming process for our applications.

Step 1: Construct Data Matrix for EachFrame

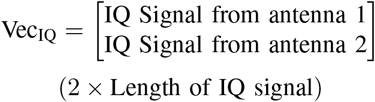

Step 2: Covariance Matrix Estimation

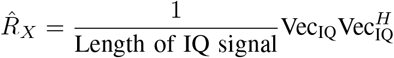

Step 3: Eigenvalue Decomposition of *R*_*X*_

- Eigenvector associated with the highest eigenvalue *A*

Step 4: Estimation of DoA

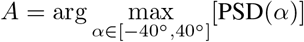

Step 5: Enhancing the SNR of the Received Signal at the Output of the Beamformer

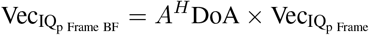

Eigen-beamforming is based on the notion of signal sub-space. In two-dimensional space (two antenna cases), the signal subspace is one dimension generated by the eigenvector associated with the highest eigenvalue. To reduce the effect of the noise (which is isotropic, i.e., occupies all dimensions) and combine coherently the two received radar reflections, we realize orthogonal projection of the steering vector on signal subspace, and we use eigenvector as a maximum power combiner to enhance Signal-to-Noise Ratio (SNR).

#### Algorithm 2 Direction of Arrival Estimation and SNR Improvement

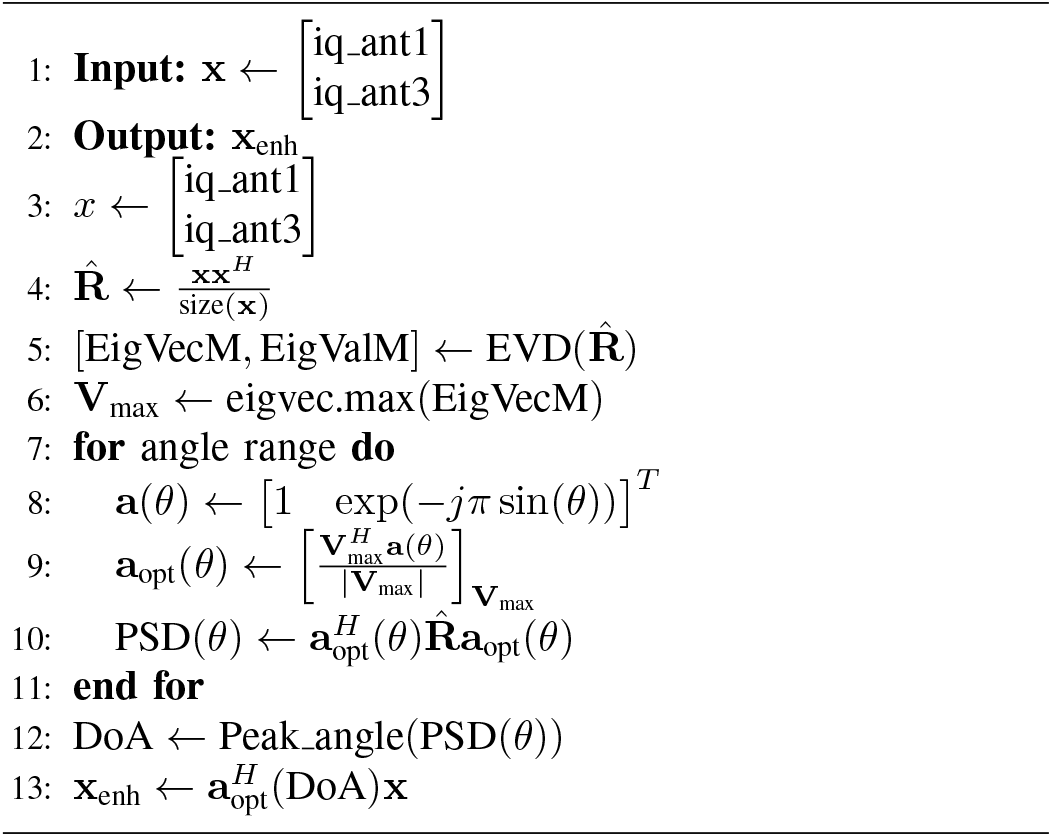

Selecting this modified version of beamforming is due to one bin selection from the FFT-range function, which induces pure sine wave function as an I/Q signal. In such one bin output signal, very low noise contribution is observed in the IQ signal, which implies an almost noise-free observation vector “*x*.” So, the covariance matrix

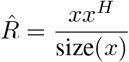

is by definition a rank-one matrix, and with almost noise-free observation, this covariance matrix is ill-conditioned (the ratio of the highest eigenvalue to the smallest eigenvalue is much greater than 1) and hence non-invertible. Constructing the steering vector (Vector of signal phases in each receiving antenna) for a given Direction of Arrival *α*:

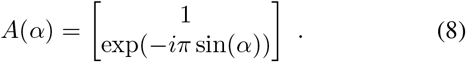

Calculating the received signal power at the output of the beamformer concerning an angle *α*:

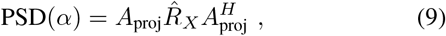

such that:

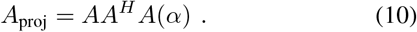

This last expression is a *A*(*α*) projection on the signal subspace generated by the eigenvector *A* associated with the highest eigenvalue. The objective of this projection is to reduce noise and other radar reflections.

### B. I/Q signal enhancement using circular regression

Since the I/Q components of radar echo create a noisy circle in the IQ plan, we have used Taubin Cicular regression to compensate for offset and noise effects in its Singular Values Decomposition (SVD) implementation to guarantee a solution, as illustrated in the Fig. 10. Taubin’s regression method was chosen for its ability to minimize algebraic errors and its robustness against noise. It typically leads to a more accurate fit than other least squares-based approaches. This technique has been effectively implemented in previous works as stated in [22] and [17].

**Fig. 10:**
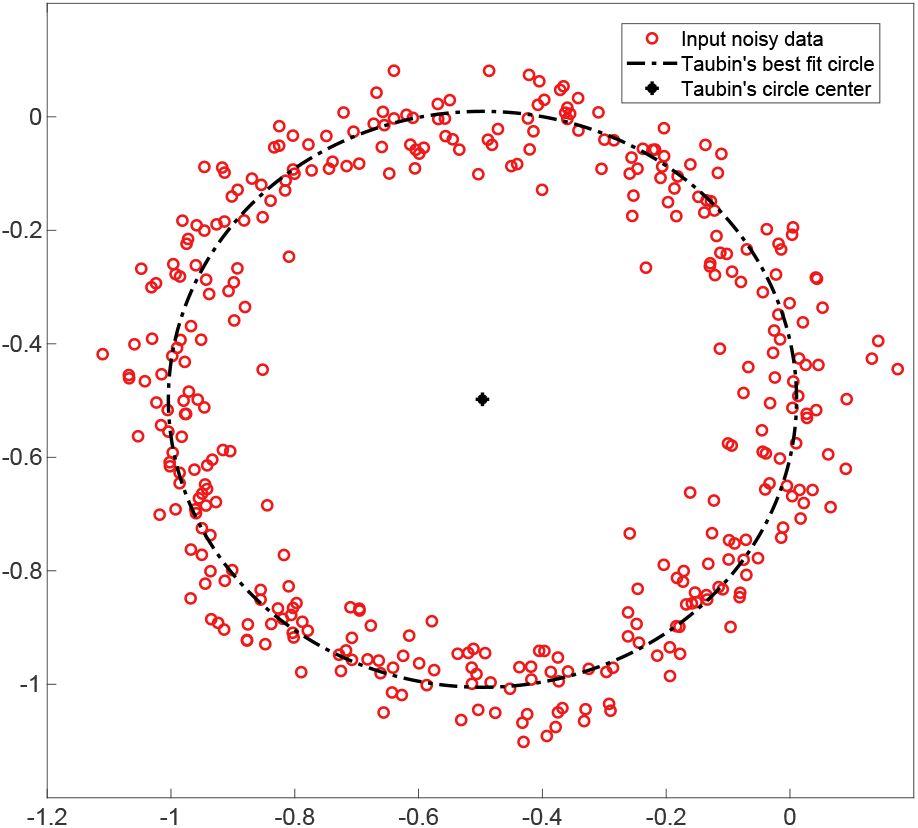
Taubin Circular Regression of Noisy and Offset Circular Data. This method minimizes algebraic distances and effectively captures the underlying circular structure in the presence of distortions.

### C. Skin displacement extraction from phase variation

To extract phase variation of reflected radar echoes caused by skin displacement, we extract the first instantaneous phase of the output signal from our eigen-beamformer. This instantaneous phase must be unwrapped to retrieve its linearly growing nature over time. Only phase fast variation rather than linear behavior of instantaneous phase carry information about jugular pulse. To extract these variations, we use phase differences between successive phase samples.

Step 1: Extracting Phase Information from IQ Enhanced Signal

- Phase estimation:

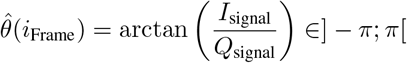
- Phase unwrapping:

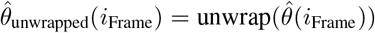
- Suppressing growing phase component:

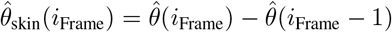

Step 2: Converting Phase Variation to Skin Displacement

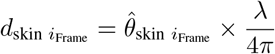

Skin displacement is related to phase variation using wave number 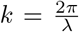 rad/m, which represents the phase variation of EM waves over space. The factor 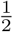 is due to the round-trip time needed for the radar signal to hit the skin and come back to its receiver.

#### Algorithm 3 Skin Displacement Estimation

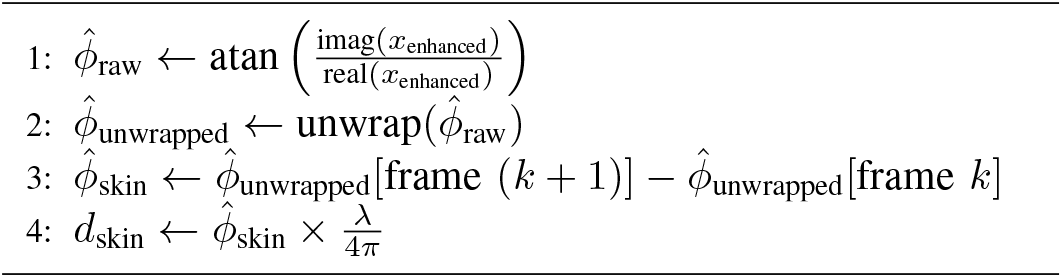

A lowpass filter (0.75–4 Hz) was applied to the displacement signal, followed by a bandstop filter (0.9–1.1 Hz) to reduce arterial pulsation impact. The filtering process was guided by a correlation threshold related to the viability of pulse waves. We select the best pulse from the recording as a reference to detect the best JVP pulses. We scan all recordings for pulses with the highest correlation coefficient using correlation. When we detect the desired number of best JVP pulses, we mark their peak values. We can choose the degree of resemblance concerning the reference pulse by fixing the correlation threshold of accepted JVP pulses. The JVP signal presents a transient effect on both ends of the recording because the filter time delay line is not fully loaded. The length of these transient portions of the signal is equal to the combined group delay of 765 samples, caused by the bandpass filter (0.75–4 Hz) and bandstop filter (0.9–1.1 Hz).

### D. SAR and PD Simulation Analysis

Specific Absorption Rate (SAR) quantifies the rate at which the body absorbs energy when exposed to an electromagnetic field, measured in watts per kilogram (W/kg). This parameter is essential for evaluating potential thermal impacts and ensuring adherence to safety regulations.

Power density refers to the amount of power distributed over a unit area perpendicular to the direction of wave propagation, indicating the intensity of power impacting a surface. Higher frequencies result in shorter wavelengths. As the wavelength decreases, the same amount of power is concentrated over a smaller area. Therefore, power density tends to increase as the frequency rises. The mechanisms and observations of these analyses can be explained in terms of skin depth and penetration. The skin depth decreases with increasing frequency, as described by the equation;

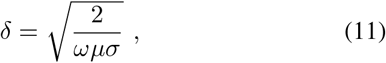

where, *ω* is the angular frequency, *μ* is the permeability, and *σ* is the conductivity of the tissue. Higher frequencies lead to smaller *δ*, meaning more energy is absorbed closer to the surface. Also, the wavelength is inversely proportional to the frequency 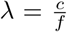, where *c* is the speed of light and *f* is the frequency.

At lower frequencies, electromagnetic waves penetrate deeper into biological tissues, resulting in a larger skin depth— where the wave power drops to about 37% of its original value—and a more dispersed energy distribution, leading to a lower SAR near the skin surface. Conversely, at higher frequencies, such as at 35 GHz, the penetration depth is much smaller, confining the wave energy to the skin’s surface layers. This concentration of energy increases the energy density and results in a higher SAR at the skin surface.

Local SAR measures the maximum energy absorbed in a small tissue volume (typically 1 g or 10 g of tissue), providing a localized exposure estimation. For the simulation, the power is averaged across the volume that encompasses 1 g of tissue using the following calculation [23];

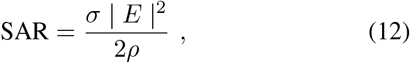

where *E* is the magnitude of the electric field, *σ* is the conductivity, and *ρ* is the mass density of biological tissue.

## VI. Results

### A. Simulation Analysis Result

The SAR analysis indicates the amount of radiation absorbed into the skin and external jugular vein, as shown in Fig. 11. At 0.9 GHz, the SAR is affected by the wave’s capacity to penetrate more deeply into the tissues, spreading the energy absorption over a larger volume and consequently yielding a lower SAR at the surface. Conversely, at 35 GHz, electromagnetic waves are absorbed at much shallower depths, concentrating more energy in the surface layers. This leads to a substantially higher SAR on the skin surface compared to lower frequencies. This simulation has enhanced our understanding of the anatomical positioning of the external jugular vein, which is very close to the skin surface.

**Fig. 11:**
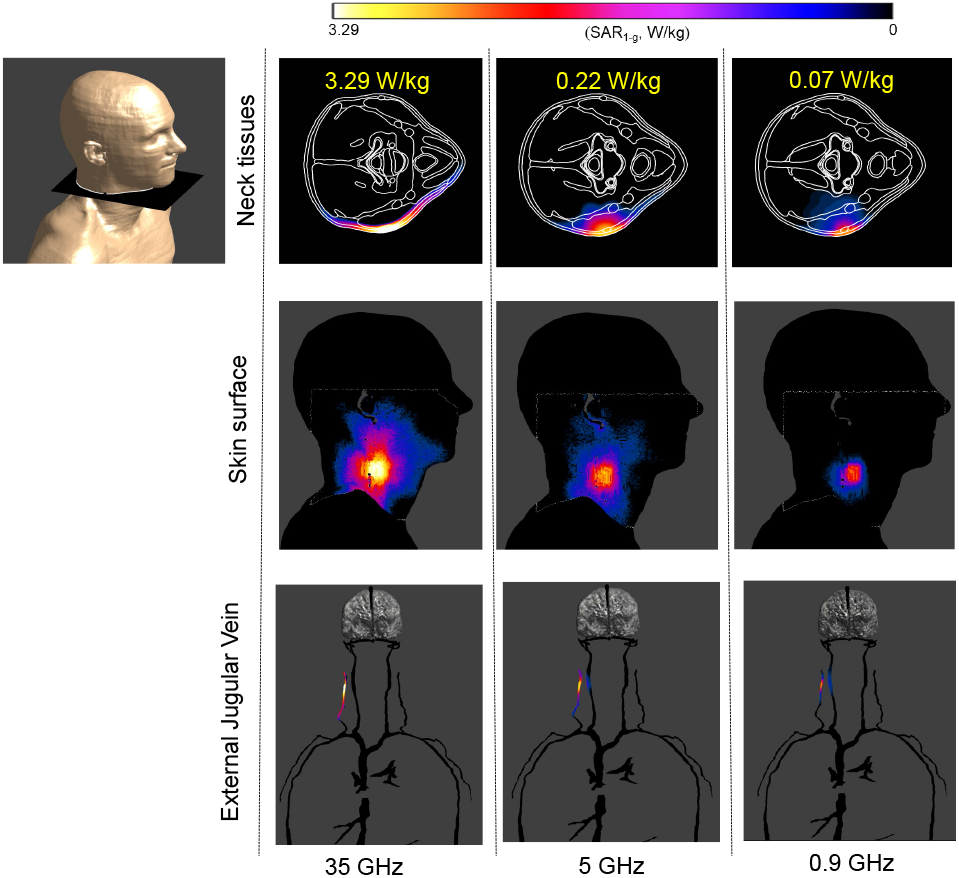
SAR analysis for skin surface, neck tissues, and the external jugular vein at frequencies of 0.9 GHz, 5 GHz, and 35 GHz. The visualization aids in understanding the electromagnetic energy absorption patterns within these tissues at various frequencies.

The SAR analysis at higher frequencies primarily targets the skin surface, with minimal absorption or scattering deeper within the tissue. Consequently, this characteristic enables a 60 GHz sensor to effectively detect the jugular pulse based on skin displacement.

The Power Density (PD) analysis of Fig. 12 indicates that as the frequency increases, the wavelength correspondingly decreases. This results in the same amount of power distributed over a smaller area. As a consequence, the power density increases with an increase in frequency. With lower frequencies, the power gets distributed in skin layers like muscle and tissue.

**Fig. 12:**
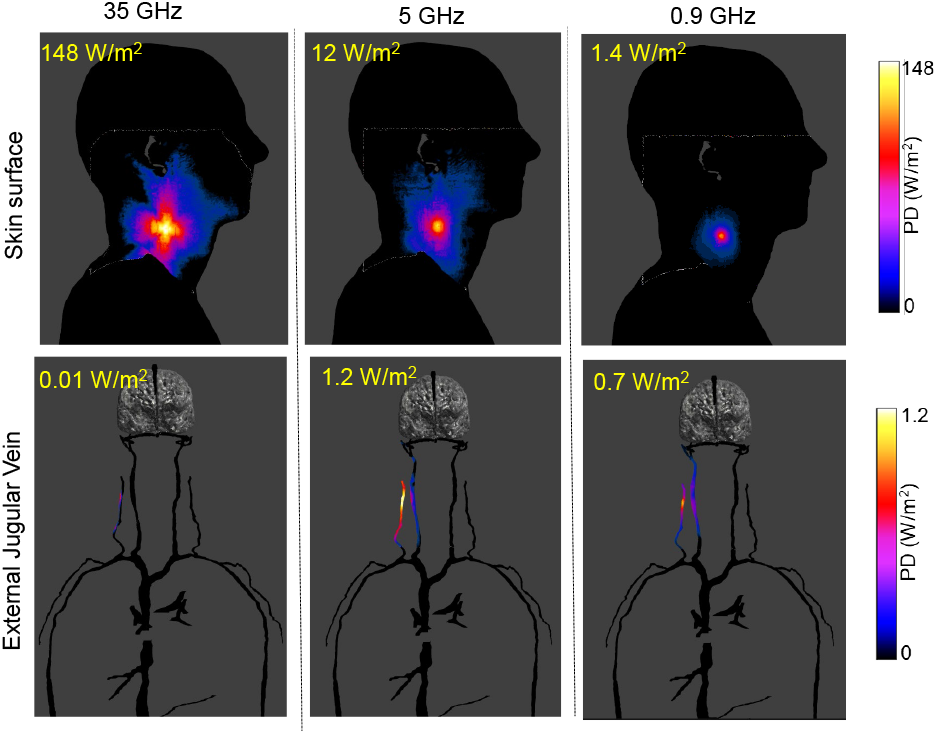
Power Density Analysis for the Skin Surface and External Jugular Vein at Frequencies of 0.9 GHz, 5 GHz, and 35 GHz. This figure displays the calculated power density values across different anatomical locations at each frequency. The results highlight significant differences in power absorption, with the highest densities observed at 35 GHz, emphasizing the frequency-dependent behavior of electromagnetic wave interactions with human tissues.

The increase in both SAR and power density with frequency is primarily due to the reduced skin depth at higher frequencies, which confines the energy absorption to the superficial layers of tissue. Understanding this frequency-dependent behavior is crucial for assessing the applications involving higher frequency ranges, such as millimeter waves. This analysis robustly demonstrates that when positioned accurately, the use of 60 GHz radar for estimating JVP predominantly captures the JVP from the skin surface, leveraging the proximity of the external jugular vein to the skin.

This detailed analysis helps elucidate the underlying physical principles governing the interaction of EM waves with biological tissues. It also highlights the importance of considering frequency when evaluating potential health risks of EM exposure.

### B. Data Analysis Result

The extraction of the JVP signal experiences amplitude fluctuations due to the non-quadrature (in-phase) radar receiver’s structure and random phase differences stemming from varying path lengths, a phenomenon validated in our acoustic experimental setup with stationary beat signals as shown in Fig. 17.

Our study explores a proof of concept technology for JVP detection using varying radar parameters, such as samples/chirp, chirps/frame, and sampling frequency. Our findings, illustrated in Fig. 13, indicate improved JVP visibility with higher samples/chirp and lower sampling frequencies; here, we used 512 samples/chirp, 8 chirps/frame, and a 500 KHz sampling frequency. The chirp duration was 132.987 *μ*s. With the controlled parameters, Fig. 14 illustrates the alteration in the JVP waveform shape when the subject torso is in an upright position. In such observation, ‘**a**’ peak was mostly visible compared to other crests.

**Fig. 13:**
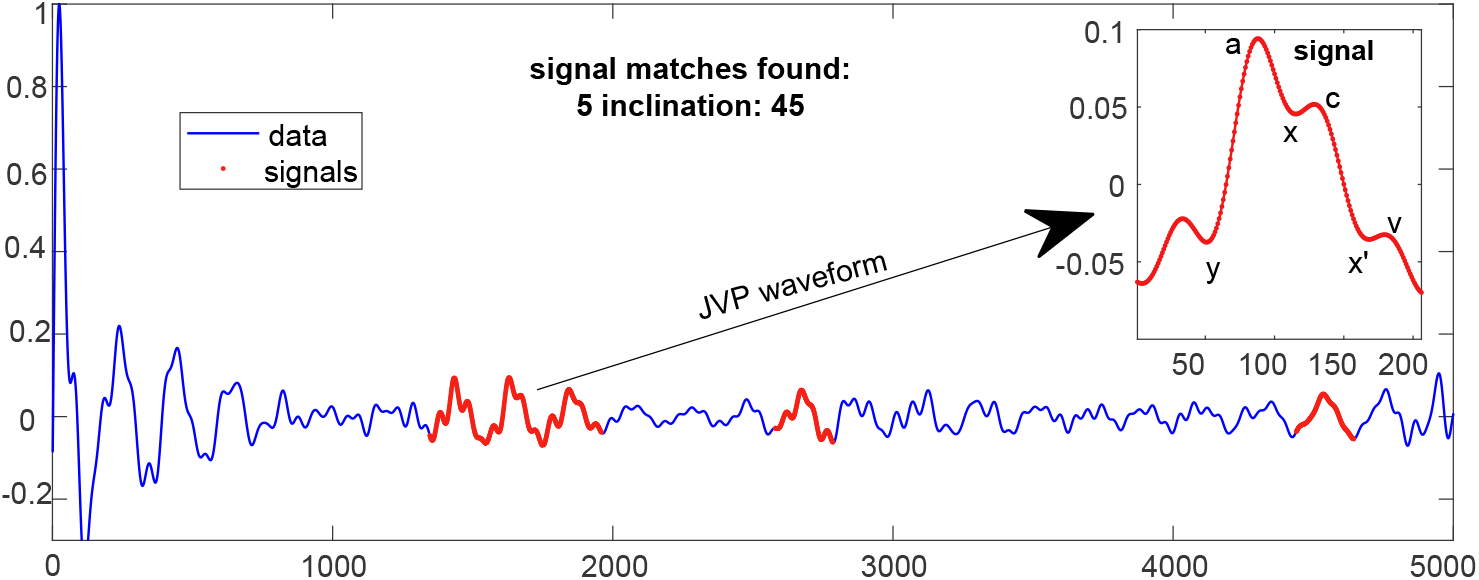
Detection of JVP waveforms in a Signal. The main graph displays the overall data signal (blue) with detected JVP waveforms highlighted in red. Signal matches found: 5. The inset (top right) zooms into a typical JVP waveform, marked from points ‘**a**’ to ‘**x**’ and ‘**x**’ to ‘**v**’ indicating the ascent and descent of the waveform, respectively.

**Fig. 14:**
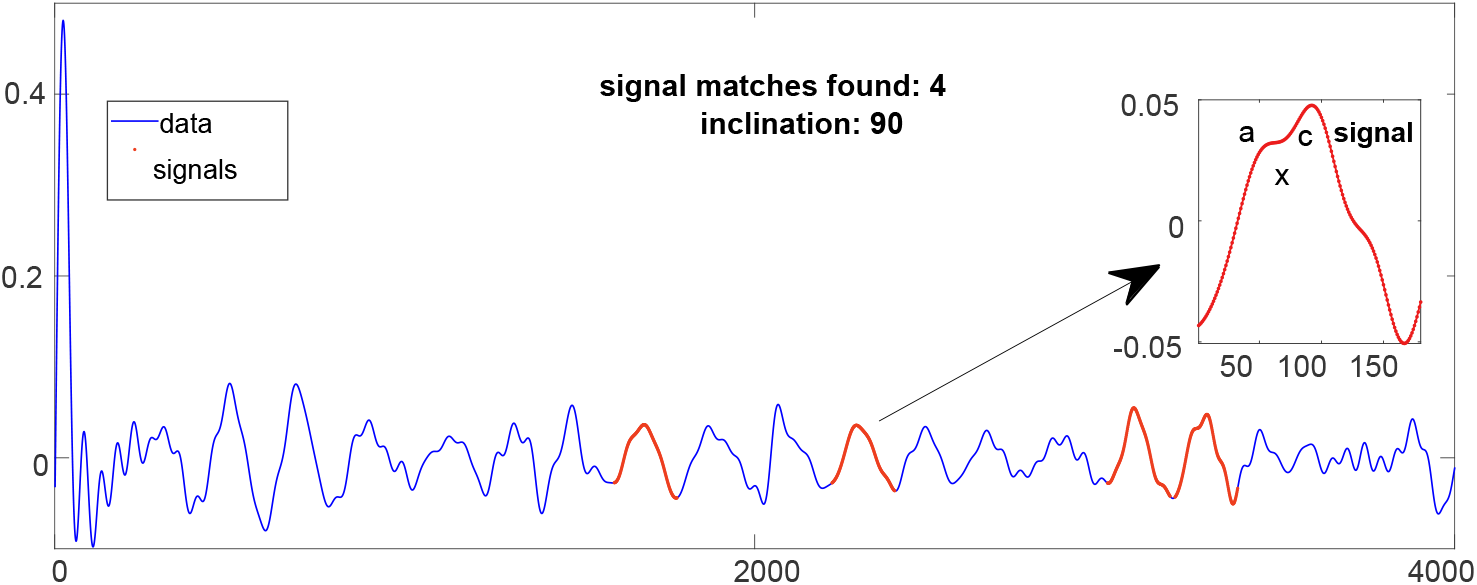
The figure shows the extracted Jugular Venous Pulse (JVP) signal at an inclination of 90 degrees. The graph depicts the signal variations over time, indicating the characteristic waveform associated with the JVP.

Observation reveals that not every pulse features distinct ‘**c**’ or ‘**v**’ peaks. Additionally, the fading phenomenon associated with this sensor has obscured the shape of some JVP signals. Hence, based on their clear morphological characteristics, we chose to show 5 of the best JVP pulses in Fig. 13. The JVP estimation hinges on pinpointing the genuine spike in the FFT. With the radar situated in proximity to the skin, the first range bin’s FFT I/Q data was leveraged to gauge skin displacement (Fig. 15(a)). Nevertheless, we also successfully extracted discernible JVP signals from the second and third bins in certain instances.

**Fig. 15:**
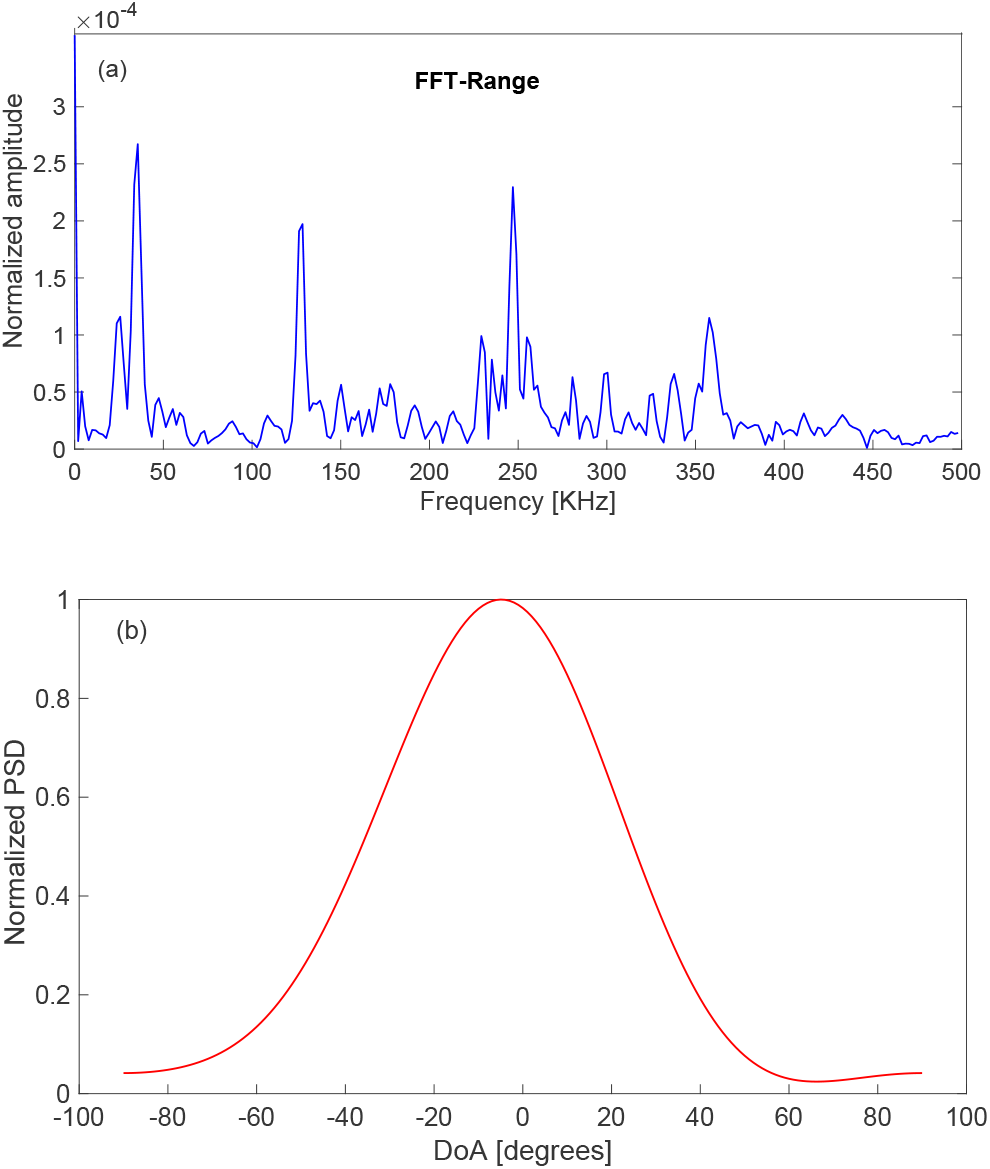
(a) FFT-Range estimation across slow time for the JVP detected at 45-degree in Fig. 13. The plot displays the normalized amplitude across various frequencies, highlighting key frequency components that characterize the JVP signal. (b) Power Spectral Density (PSD) distribution for DoA estimation using radar sensor. The graph shows the normalized PSD across different angles, revealing a prominent peak around *−*5^°^, indicating the estimated direction of the signal source.

Skin displacement measurement effectiveness can differ based on each antenna’s signal quality. Due to challenges with ill-conditioned covariance matrices in the beamforming algorithm from [17], we exclusively adopted the previously mentioned eigen-beamformer method, achieving a similar SNR as both beamforming approaches produce co-linear vectors.

The DoA refers to the angle at which a received signal arrives relative to a reference point or axis. In radar systems used for physiological monitoring, when the body inclines or changes position, the geometry of signal reflection alters. This means the reflected signals may travel different paths, changing the perceived DoA at the radar sensor. The DoA estimation for JVP ranges between −20^°^ and 20^°^ for different inclinations. Fig. 15(b) illustrates the DoA estimation of JVP waveform when tilted at a steep angle.

Fig. 16 shows the representation when the upper body of the subject was upright with the PPG recorded simultaneously from the right index finger using the mentioned device. A time delay of approximately 1.2884 seconds was noted, likely influenced by equipment buffering, processing delays, and frequency variations. Conventionally, the JVP valley should match the PPG signal’s peak. Here, we can observe this match at 4, 5, 6, 11 and 15 secs. It is known that due to gravitational influence and reduced hydrostatic pressure in the vein, the visibility of JVP at 90^°^ is very lower, but we managed to get a visible JVP when upright by sticking to our controlled parameters of the radar.

**Fig. 16:**
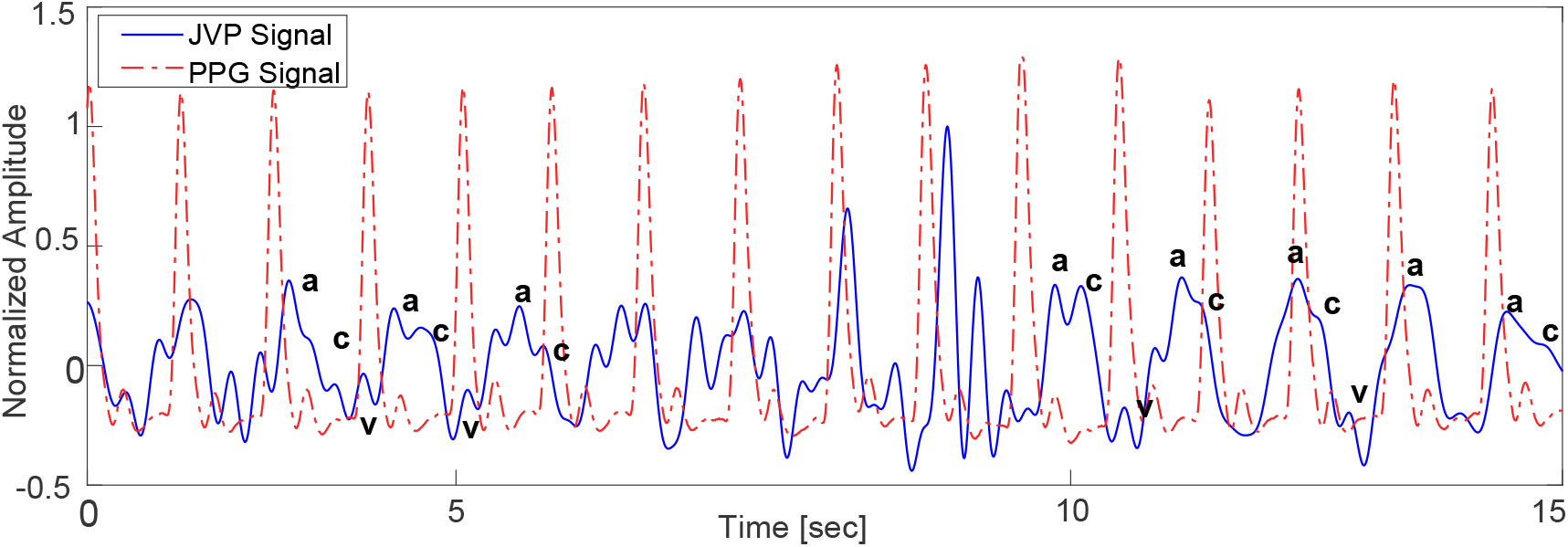
Comparative analysis of manually annotated ‘**a**’ and ‘**c**’ peaks of the JVP alongside the reference PPG signal obtained from a finger at a 90^°^ angle. This illustration highlights the temporal alignment and amplitude differences between the venous pulsations observed in the neck and the arterial pulsations measured peripherally.

Fig. 17 shows the estimated JVP for different parameters when the sampling frequency is higher at 1 GHz with samples per chirp as 128 and chirps per sample as 8. Though we can visualize some JVPs, not all of them are of the same quality. These findings enhance our comprehension of parameter adjustments necessary for improved JVP visibility.

**Fig. 17:**
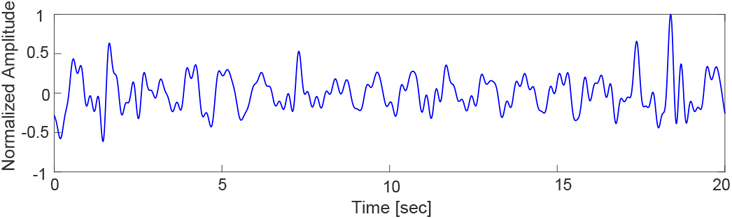
JVP signal acquired at an angle of 45^°^. The data was sampled at a high frequency of 1 GHz, capturing 128 samples per chirp over a sequence of 8 chirps.

## VII. Discussion

Our experiment clearly demonstrates that the 60 GHz FMCW radar effectively performs non-invasive extraction of JVP. The selection of a 60 GHz radar system is attributed to its superior resolution and compact design featuring integrated antennas, which enhances its portability. Also, its high sensitivity is crucial for detecting subtle changes in the skin’s surface caused by the underlying venous pulsations. In our scenario involving proximity measurements, we employed the maximum bandwidth of 5.5 GHz for all recordings.

It became apparent that several factors influence JVP performance, notably the distance between the radar and the subject. A radar positioned too closely can cause the initial spike to blend with the DC component in the FFT, complicating the identification of the actual spike. Accurately detecting this spike is crucial for achieving high-resolution JVP estimations related to skin displacement. On the contrary, if the radar is too far, it cannot detect the pulse as the amplitude of JVP is very low. Therefore, we tried to keep an optimum distance of 2 mm from the skin surface for data recording.

Further observation reveals that reciprocal mixing among FFT-range bins mandates adjustments to the radar parameters and experimental setup enhancements for optimal JVP quality. Despite the radar’s capability for parameter customization, the hardware imposes a strict correlation between the sampling frequency and the number of samples per chirp. The utilized Infineon’s radar functions through fast chirp sequences, presenting a challenge in detecting phase changes with just two chirps due to their rapid generation of chirps. To effectively extract phase information from the radar signals, a minimum of 8 chirps is essential for achieving accurate results. Through meticulous evaluation, we established the subsequent configuration for JVP estimation with this radar:

- Higher sampling rate for a fast time (higher number of samples per chirp) to increase FFT-range resolution. Because a lower value implies a reduction of the signal quality and even burying the signal in noise;
- Suppress averaging on FFT-range spectrums over all chirps to avoid losing JVP signal details;
- Higher frame rate to get much more time resolution for JVP signal.
- In some situations, a signal with low SNR in one antenna can significantly reduce the performance of the beamformer and, eventually, the pulse detection. Therefore, an antenna selection procedure must be adopted;

Also, a shorter chirp duration is helpful because if the chirp is longer, the chirp reflection collides with the transmitted one. So, for better results, we narrowed down the parameter values as the following:

**TABLE I:**
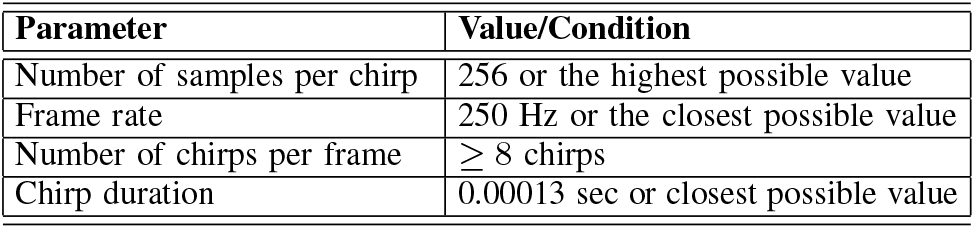
Radar Configuration for optimal JVP estimation.

| Parameter | Value/Condition |
| --- | --- |
| Number of samples per chirp | 256 or the highest possible value |
| Frame rate | 250 Hz or the closest possible value |
| Number of chirps per frame | $\geq 8$ chirps |
| Chirp duration | 0.00013 sec or closest possible value |

### Note

Longer chirp durations are permissible if the radar-skin distance increases. Additionally, increasing the receiver’s gain is essential under these conditions.

The chip receiver incorporates a single in-phase mixer, lacking a quadrature (Q) component, and thus generates a real-valued IF signal. A Fourier transform is applied to this IF signal during the processing, resulting in complex I and Q signals that carry phase information for each frequency component. An accurate radar provides time-varying performance compared to IQ radar due to phase variation. This architecture suffers from a fading phenomenon due to phase offset between the reference signal and radar reflection. As illustrated in the following figure, the signal is a function of time. Through the configuration illustrated in the figure simulator Fig. 18, we establish the critical enhancements offered by quadrature (Q) radar receivers over non-quadrature or in-plain (I) counterparts, as outlined in [24].

**Fig. 18:**
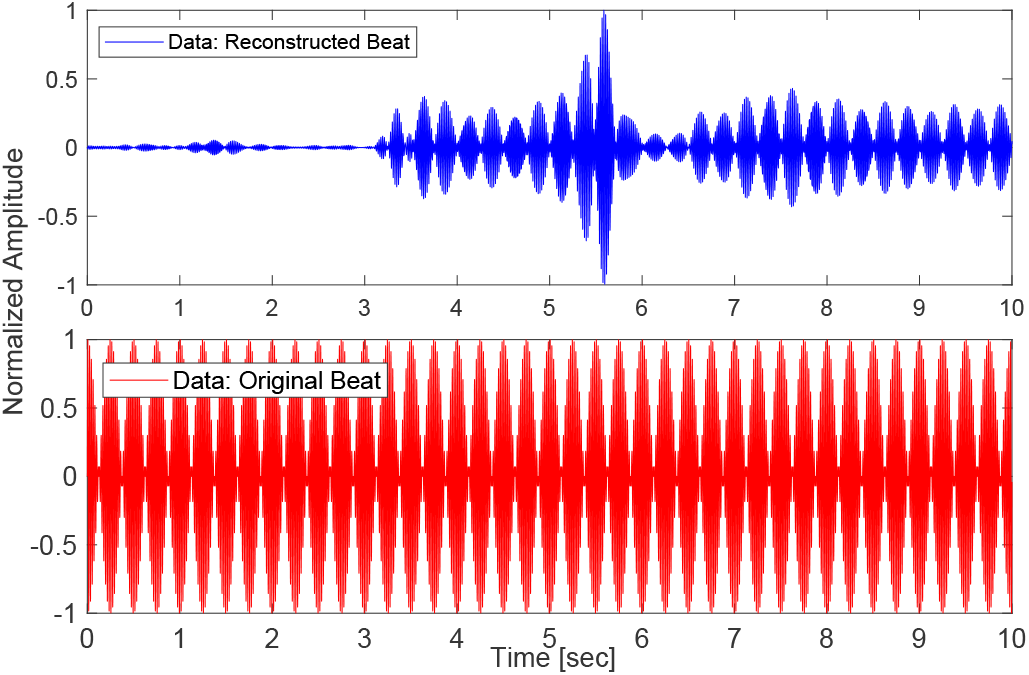
(Top): Beat signal reconstructed by Radar, (Bottom): Original signal played on speaker

The original beat signal exhibits a pronounced spike thanks to its high amplitude, while radar-based beat reconstruction reveals an inconsistent envelope signal stemming from the phase difference between the received signal and the local oscillator. This experiment not only confirms the effectiveness of our algorithm but also sheds light on the constraints related to the hardware. Therefore, we prefer to use a sensor equipped with an IQ mixer for future experiments. In the phase extraction process, we employed the Taubin Circular Regression method to identify the optimally centered circle that aligns with our IQ signal, thereby compensating for any offset and distortion. Furthermore, the selection of the eigen-beamformer played a crucial role in addressing the issue of the poorly conditioned correlation matrix.

Often, JVP signals can overlap with or are superimposed by stronger signals from arterial pulsations or respiratory movements. This overlap can mask the delicate JVP waveform, making it harder to isolate and analyze. Although [6] explored non-contact JVP measurement with a 24 GHz microwave radar, it did not account for the influence of arterial signals. Recognizing this challenge in our research, we employed a supplementary bandstop filter specifically designed to diminish the influence of arterial pulsations on the JVP signal. Our DoA measurement gives us an accurate understanding of the localization of the JVP signal.

Inspired by the application of 60 GHz radar for blood pressure estimation in [17], we employed a radar to explore and analyze JVP as a proof of concept. Our study demonstrated the radar’s potential in JVP estimation, overcoming its design limitations.

## VIII. Ethics Statement

The ethics considerations for this study are presented to, reviewed, and approved by the Human Research Ethics Committee (HREC) at The University of Sydney under [2024/104].

## IX. Conclusion

This study presents a proof of concept using an FMCW radar sensor at 60 GHz for non-invasive JVP estimation, offering a cost-effective alternative to traditional methods. Despite encountering certain hardware and design limitations, we successfully measured the nuanced, millimeter-scale JVP and identified optimal parameters for its estimation with this technology. This sensor demonstrated its value in precisely determining the DoA. We also conducted a comparative analysis with simultaneously recorded finger PPG and explored the impact of various body inclinations on JVP measurements. It’s important to recognize that physiological factors such as body mass index and the composition of fat and tissue can influence JVP readings, given the jugular vein’s location beneath muscle, skin, and fat layers. Future research will expand on these findings in a larger cohort. In summary, the 60 GHz FMCW radar has proven its potential in JVP estimation and offers promising insights. Addressing the current challenges could lead to an effective tool for early right HF detection, significantly reducing hospital admissions by providing a valuable resource for clinicians and patients, especially in remote settings.

## Data Availability

All data produced in the present study are available upon reasonable request to the authors

## Footnotes

1 For further details, please contact the corresponding author at

## Notes

This project is supported by The Medical Research Future Fund (MRFF) Cardiovascular Health Mission grant (number: MRF2017760). The authors acknowledge using ZMT Zurich MedTech AG’s Sim4Life, Zurich, Switzerland, in this work’s electromagnetic simulations.

### Competing Interest Statement

The authors have declared no competing interest.

### Funding Statement

This project is supported by The Medical Research Future Fund
(MRFF) Cardiovascular Health Mission grant (number: MRF2017760)

